# *Staphylococcus aureus* Tetracycline Resistance and Co-resistance in a Doxy-PEP-Eligible Population

**DOI:** 10.1101/2024.08.22.24312434

**Authors:** Rachel Mittelstaedt, Sanjat Kanjilal, David Helekal, Gregory K. Robbins, Yonatan H. Grad

## Abstract

In *Staphylococcus aureus* infections in men eligible for doxycycline post-exposure prophylaxis (doxy-PEP), tetracycline non-susceptibility is more prevalent than in the overall population and is associated with resistance to trimethoprim-sulfamethoxazole and clindamycin. Doxy-PEP may select for *S. aureus* multi-drug resistance, underscoring the importance of surveillance.

## Introduction

Rates of bacterial sexually transmitted infections (STIs) are increasing in the United States.^1^ Several randomized control trials demonstrated that doxycycline post-exposure prophylaxis (doxy-PEP), prescribed as 200 mg of doxycycline taken within 72 hours of condomless sex, reduces the incidence of bacterial STIs among men who have sex with men (MSM) and transgender women (TGW).^2^ These findings prompted national guidelines recommending consideration of doxy-PEP for MSM and TGW, particularly those who have had a bacterial STI in the past twelve months.^3^

One concern with doxy-PEP is that it may select for resistance to tetracyclines and other antibiotics both in the targeted pathogens and in bystander colonizing bacteria that have pathogenic potential. Supporting this concern, daily use of doxycycline selected for increased tetracycline resistance in the skin microbiome.^4^ Additionally, the US-based DoxyPEP study found increased rates of *Staphylococcus aureus* doxycycline resistance among participants receiving doxy-PEP, though the results were not statistically significant in the setting of a small sample size.^2^

While increasing rates of tetracycline resistance could create challenges in the treatment of many bacterial pathogens^5^, resistance in *S. aureus* is of particular concern. Doxycycline is a preferred therapy for stepdown and outpatient management of *S. aureus*, including methicillin resistant *Staphylococcus aureus* (MRSA).^6^ Furthermore, because resistance tends to aggregate in certain strains, selection of tetracycline-resistant *S. aureus* by doxy-PEP could lead to co-selection for strains that are resistant to other anti-staphylococcal antibiotics.^7^ Here, we sought to characterize patterns of *S. aureus* resistance and co-resistance in patients who may be prescribed doxy-PEP in an effort to understand the potential impact of widespread doxy-PEP adoption.

## Methods

We performed a retrospective analysis of all *S. aureus* cultures in people with a sex of male reported in the electronic medical record (EMR) who were either HIV-positive or prescribed HIV pre-exposure prophylaxis (PrEP), 18 years of age or older, and receiving care at two academic medical centers in Boston, MA between June 2015 and May 2022. Patients were identified using a clinical data repository containing microbiologic data, diagnosis codes, and medication prescriptions from the EMR. Because the number and gender of sexual partners was not reliably recorded in the EMR, HIV positivity and receipt of HIV-PrEP were used to identify patients whose sexual behaviors were most likely to qualify them for doxy-PEP under current guidelines.^3^

Unique episodes of infection were defined as all cultures that were positive for *S. aureus* with identical antibiotic susceptibility profiles within a 14-day period. Isolates were considered distinct if their susceptibility to any antibiotic was ≥ two 2-fold dilutions removed from the minimum inhibitory concentration (MIC) of the first isolated strain. If multiple *S. aureus* specimens with distinct susceptibility profiles were isolated during a given 14-day period, all were included (**Table S1**). Nasal swabs performed for MRSA surveillance were not included because they were not assessed for tetracycline susceptibility.

Susceptibility of *S. aureus* isolates to tetracycline, doxycycline, oxacillin, trimethoprim-sulfamethoxazole (TMP/SMX), and clindamycin were defined by Clinical and Laboratory Standards Institute (CLSI) MIC breakpoints.^8^ Isolates with inducible resistance to clindamycin were categorized as resistant. The frequency of resistance to oxacillin, TMP/SMX, and clindamycin in tetracycline-susceptible *S. aureus* isolates (TET-S, MIC ≤ 4.0 μg/mL) was compared to the frequency of resistance in the same antibiotics among tetracycline intermediate (TET-I, MIC = 8 μg/mL) or tetracycline resistant isolates (TET-R, MIC ≥ 16 μg/mL). Rates of institutional resistance were calculated using institution-specific antibiograms from 2022 which reported the resistance patterns of 5988 total isolates (2300 from the Brigham and Women’s Hospital and 3477 from the Massachusetts General Hospital). Fisher’s exact test was used to assess resistance co-occurrence and adjusted for multiple comparisons using the Bonferroni correction. Logistic regression was used to assess for association between year of sampling and resistance over the study period. Statistical tests were performed in R (version 4.1.2, R Core Team 2021),^9^ and the *tidyverse* package was used for data analysis and visualization.^10^

## Results

We identified 832 isolates of *S. aureus* collected from 543 patients. The mean number of isolates collected per patient was 1.53, with a median of 1 and a maximum of 16 (**Table S1**). Most isolates were obtained from a skin/soft tissue source (280, 33.7%), an abscess (162, 19.5%), the lungs (126, 15.1%), or blood (83, 10.0%) (**Table S2**). While there was no statistically significant association between resistance to doxycycline, clindamycin, and TMP/SMX and the year of sampling, there was a statistically significant association between the year of sampling and resistance to tetracycline (p = 0.002) and oxacillin (p = 0.002), both of which increased over the study period (**Figure S1**). Isolates from our patient cohort were significantly more resistant to oxacillin, TMP/SMX, and clindamycin than the institutional averages reported in 2022 (**Tables S3 and S4**).

Of the 832 isolates, 829 had tetracycline susceptibility data, and 620 had doxycycline susceptibility data. Among isolates with tetracycline susceptibility data, 116 isolates were tetracycline resistant (MIC ≥ 16 μg/mL) and 4 isolates were tetracycline intermediate (MIC = 8 μg/mL). Among isolates with doxycycline susceptibility data, 10 were doxycycline resistant (MIC ≥ 16 μg/mL) and 18 were doxycycline intermediate (MIC = 8 μg/mL).

Patterns of co-resistance associated with tetracycline non-susceptibility are summarized in **Table 1**. There was no statistically significant association between tetracycline non-susceptibility and oxacillin resistance (OR 1.35, 95% CI 0.83 – 2.21, p = 0.42). However, tetracycline non-susceptible isolates were 2.32 times more likely to be resistant to TMP/SMX (95% CI 1.20 – 4.32, p = 0.004), and 2.33 times more likely to be resistant to clindamycin (95% CI 1.43 – 3.82, p < 0.001) than tetracycline-susceptible isolates.

**Table 1:** Odds of resistance to oxacillin, trimethoprim-sulfamethoxazole, and clindamycin in the setting of tetracycline non-susceptibility. Counts and percentages of *S. aureus* isolates that are susceptible to tetracycline (MIC ≤ 4.0 μg/mL) or intermediate/resistant to tetracycline (MIC ≥ 8.0 μg/mL) and susceptible (MIC ≤ 2.0 μg/mL) or resistant (MIC ≥ 4.0 μg/mL) to oxacillin, susceptible (MIC ≤ 2/38 μg/mL) or resistant (MIC ≥ 4/76) to trimethoprim-sulfamethoxazole, or susceptible (MIC ≤ 0.5 μg/mL without inducible resistance) or resistant (MIC ≥ 4 μg/mL or positive for inducible resistance) to clindamycin. Odds ratios (OR) of co-resistance to oxacillin, trimethoprim-sulfamethoxazole, and clindamycin in the setting of tetracycline non-susceptibility are reported. There was a significant association between tetracycline non-susceptibility and resistance to TMP/SMX and clindamycin. TET = tetracycline; OXA = oxacillin; TMP/SMX = trimethoprim-sulfamethoxazole; CLI = clindamycin.

| Oxacillin |  |  |  |
| --- | --- | --- | --- |
|  | OXA-Resistant | OXA-Sensitive | OR (CI, p-value) |
| TET-Intermediate/Resistant | 63 (51.6%) | 59 (48.4%) | 1.35 (0.83 – 2.21, p = 0.42) |
| TET-Sensitive | 311 (44.1%) | 394 (55.9%) |  |
| Trimethoprim-Sulfamethoxazole |  |  |  |
|  | TMP/SMX-Resistant | TMP/SMX-Sensitive | OR (CI, p-value) |
| TET-Intermediate/Resistant | 26 (21.1%) | 97 (78.8%) | 2.32 (1.20 – 4.32, p = 0.004) |
| TET-Sensitive | 73 (10.3%) | 633 (89.7%) |  |
| Clindamycin |  |  |  |
|  | CLI-Resistant | CLI-Sensitive | OR (CI, p-value) |
| TET-Intermediate/Resistant | 63 (51.2%) | 60 (48.8%) | 2.33 (1.43 – 3.82, p <0.001) |
| TET-Sensitive | 219 (31.0%) | 487 (69.0%) |  |

We assessed rates of resistance to doxycycline in tetracycline-susceptible and tetracycline-intermediate/resistant isolates. There were 620 isolates with both doxycycline and tetracycline MICs, with 524 tetracycline-susceptible and 96 tetracycline-intermediate/resistant isolates. Among tetracycline-susceptible isolates, 524 (100%) were also susceptible to doxycycline. Among tetracycline-intermediate/resistant isolates, 68 were susceptible (70.6%), 18 intermediate (18.8%), and 10 resistant (10.4%) to doxycycline (**Figure S2**).

## Discussion

High-risk sexual practices are a risk factor for community-acquired MRSA in MSM living with HIV.^11^ In keeping with this finding, *S. aureus* infections in our cohort of doxy-PEP eligible patients demonstrated increased rates of resistance to commonly used anti-staphylococcal antibiotics when compared to the institutional average. Furthermore, we found that tetracycline non-susceptibility in *S. aureus* was significantly associated with co-resistance to two anti-staphylococcal antibiotics, TMP/SMX and clindamycin, in a cohort of patients with a sex of male reported in the EMR who were either HIV-positive or prescribed HIV PrEP. While doxy-PEP offers a promising strategy to decrease the frequency of bacterial STIs, increased selective pressure from higher rates of doxycycline use^12^ could foster tetracycline resistance in *S. aureus* and other common pathogens via bystander selection. Our results suggest that in selecting for tetracycline resistance, doxy-PEP could select for strains of *S. aureus* that are also resistant to TMP/SMX and clindamycin.

Limitations to this study include using HIV positivity and PrEP as a proxy for identifying those who might be prescribed doxy-PEP. This strategy likely excluded some who might be doxy-PEP eligible, such as MSM and TGW who are HIV-negative and not on PrEP, and included others in whom doxy-PEP is not currently recommended, such as men who have sex with women. Additionally, we used tetracycline susceptibility as a proxy for susceptibility to doxycycline, following CLSI guidelines.^9^ However, many tetracycline-resistant isolates were reported as doxycycline-susceptible. Our cohort included too few examples of isolates with doxycycline MICs greater than ≥16 μg/mL to draw meaningful conclusions about co-inheritance of resistance to other antibiotics at this threshold. As the strength of doxy-PEP selection for tetracycline resistance in *S. aureus* is unknown, we made the conservative choice to evaluate at the CLSI cutoff for tetracycline non-susceptibility.

To date, studies examining development of antimicrobial resistance in *S. aureus* after use of doxy-PEP have been limited in size and focused on resistance to doxycycline. Our findings suggest that the doxy-PEP eligible population is already more likely to carry resistant strains of *S. aureus*. Furthermore, use of doxy-PEP in this population may select for strains of *S. aureus* that carry resistance to not only tetracyclines, but also to other common anti-staphylococcal antibiotics. The extent to which doxy-PEP will select for these strains and the efficiency with which resistant strains will be transmitted is unknown. Thus, it will be important to consider monitoring *S. aureus* resistance to tetracyclines and to other classes of antibiotics in patients taking doxy-PEP.

## Data Availability

The data used in the present study is protected health information and therefore not publicly available. Upon request to the authors, a de-identified sample dataset can be provided.

## Acknowledgements

The authors thank Kirstin Roster and Thi Mui Pham for helpful comments on the manuscript. The project described was supported by Grant Number T32 AI007433 from the National Institute of Allergy and Infectious Diseases. Its contents are solely the responsibility of the authors and do not necessarily represent the official views of the NIH.

This project was funded in part by contract 200-2016-91779 with the Centers for Disease Control (CDC) and Prevention. Disclaimer: The findings, conclusions, and views expressed are those of the authors and do not necessarily represent the official position of the CDC.

## References

1. Centers for Disease Control and Prevention. Sexually Transmitted Disease Surveillance 2022. Atlanta: U.S. Department of Health and Human Services; 2024.

2. Luetkemeyer AF, Donnell D, Dombrowski JC, et al. Postexposure Doxycycline to Prevent Bacterial Sexually Transmitted Infections. N Engl J Med. 2023;388(14):1296–1306. doi:10.1056/NEJMoa2211934

3. Bachmann LH, Barbee LA, Chan P, et al. CDC Clinical Guidelines on the Use of Doxycycline Postexposure Prophylaxis for Bacterial Sexually Transmitted Infection Prevention, United States, 2024. MMWR Recomm Rep. 2024;73(2):1–8. doi:10.15585/mmwr.rr7302a1

4. Jo JH, Harkins CP, Schwardt NH, et al. Alterations of human skin microbiome and expansion of antimicrobial resistance after systemic antibiotics. Sci Transl Med. 2021;13(625):eabd8077. doi:10.1126/scitranslmed.abd8077

5. Kenyon C. Doxycycline post-exposure prophylaxis could theoretically select for resistance to various antimicrobials in 19 pathobionts: an in silico analysis. International Journal of Infectious Diseases. 2024;142:106974. doi:10.1016/j.ijid.2024.02.017

6. Liu C, Bayer A, Cosgrove SE, et al. Clinical Practice Guidelines by the Infectious Diseases Society of America for the Treatment of Methicillin-Resistant Staphylococcus aureus Infections in Adults and Children. Clinical Infectious Diseases. 2011;52(3):e18–e55. doi:10.1093/cid/ciq146

7. Chang HH, Cohen T, Grad YH, Hanage WP, O’Brien TF, Lipsitch M. Origin and Proliferation of Multiple-Drug Resistance in Bacterial Pathogens. Microbiol Mol Biol Rev. 2015;79(1):101–116. doi:10.1128/MMBR.00039-14

8. CLSI. Performance Standards for Antimicrobial Susceptibility Testing. 34th ed. CLSI supplement M100. Clinical and Laboratory Standards Institute; 2024.

9. R Core Team (2021). R: A language and environment for statistical computing. R Foundation for Statistical Computing, Vienna, Austria. URL https://www.R-project.org/.

10. Wickham H, Averick M, Bryan J, Chang W, McGowan LD, François R, Grolemund G, Hayes A, Henry L, Hester J, Kuhn M, Pedersen TL, Miller E, Bache SM, Müller K, Ooms J, Robinson D, Seidel DP, Spinu V, Takahashi K, Vaughan D, Wilke C, Woo K, Yutani H (2019). “Welcome to the tidyverse.” _Journal of Open Source Software_, *4*(43), 1686. doi: 10.21105/joss.01686 (URL: https://doi.org/10.21105/joss.01686).

11. Lee NE, Taylor MM, Bancroft E, et al. Risk Factors for Community-Associated Methicillin-Resistant Staphylococcus aureus Skin Infections among HIV-Positive Men Who Have Sex with Men. Clin Infect Dis. 2005;40(10):1529–1534. doi:10.1086/429827

12. Roster KIO, Grad YH. Estimating Changes in Antibiotic Consumption with the Introduction of Doxycycline Post-Exposure Prophylaxis in the United States. Public and Global Health; 2023. doi:10.1101/2023.09.20.23295787

