## Supplement for "*Staphylococcus aureus* Tetracycline Resistance and Co-resistance in a Doxy-PEP-Eligible Population"

**Supplemental Material**

**Figure S1**: **Resistance Trends in *S. aureus* from 2015 – 2022**
The percentage of *S. aureus* isolates in our cohort resistant to clindamycin, doxycycline, oxacillin, tetracycline, and TMP/SMX between June 1, 2015, and May 2, 2022. TMP/SMX = trimethoprim-sulfamethoxazole. This figure was prepared in R (version 4.1.2, R Core Team 2021)[1] using the *tidyverse*[2] and *jtools*[3] packages.

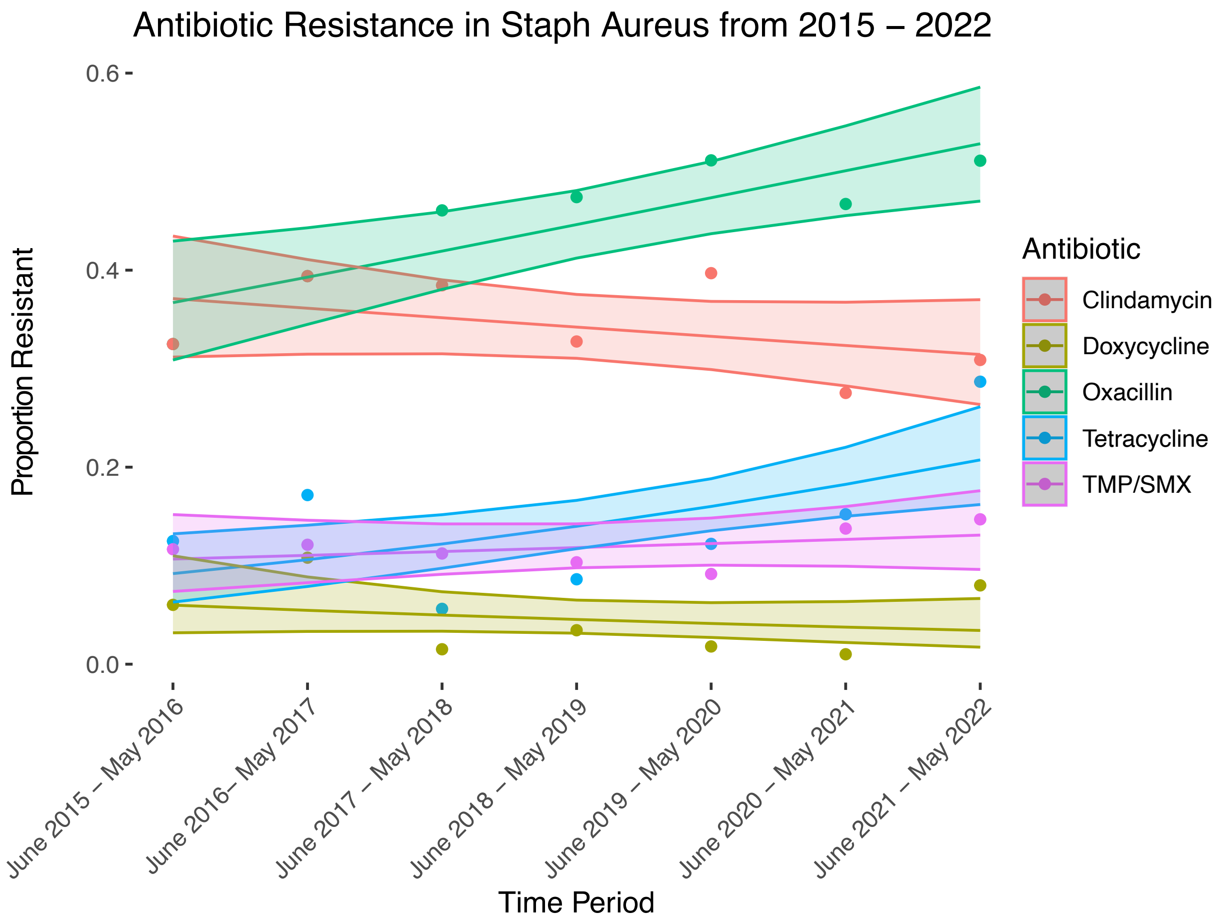

**Figure S2: Correlation of Tetracycline Resistance to Doxycycline Resistance**
**A.** Counts and percentages of *S. aureus* isolates that are susceptible to tetracycline (TET-S, MIC ≤ 4.0 µg/mL) or intermediate/resistant to tetracycline (TET-IR, MIC ≥ 8.0 µg/mL) and susceptible (MIC ≤ 4.0 µg/mL), intermediate (MIC = 8), or resistant (MIC ≥ 16 µg/mL) to doxycycline. **B.** A magnified view of counts and percentages of tetracycline intermediate/resistant isolates by doxycycline minimum inhibitory concentrations. All isolates described in Panel B have a tetracycline MIC ≥ 8.0 µg/mL. Only 29.2% of tetracycline non-susceptible isolates were also doxycycline non-susceptible. This figure was prepared in R (version 4.1.2, R Core Team 2021)[1] using the *tidyverse*[2] package.

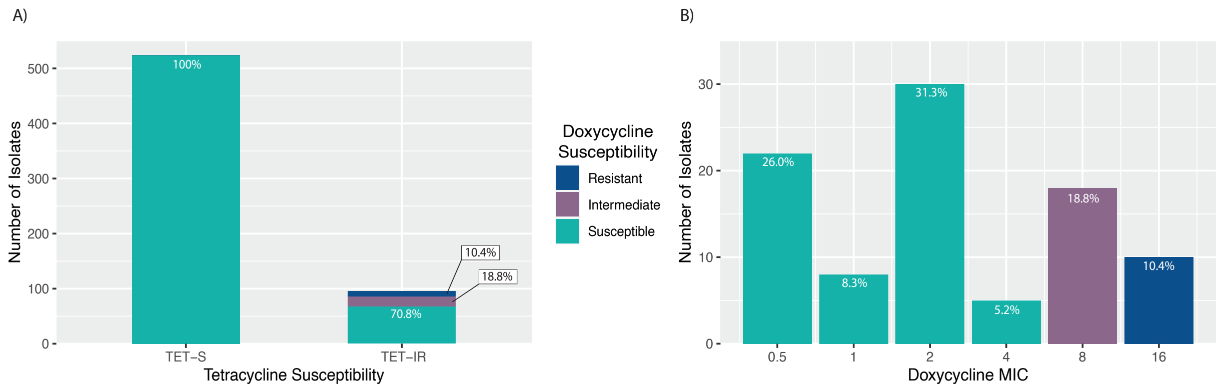

**Table S1: Number of Specimens Contributed per Patient**

This table describes the number of isolates contributed per patient. 543 total patients contributed 832 total specimens, with a mean number of 1.53 isolates per patients. 390 patients contributed 1 specimen, 87 patients contributed 2 specimens, 40 patients contributed 3 specimens, etc. Clinical records of patients contributing more than 10 specimens were manually reviewed. In two cases, review indicated patients with 20 specimens had chronic infection or colonization with *S. aureus*, in which the specimens did not reflect distinct episodes of acute infection. In these cases, only unique strains of *S. aureus* were included in the final analysis, reducing the total number of specimens from these patients to 6 each.

| Number of Specimens Contributed per Patient | Number of Patients |
|---|---|
| 1 | 390 |
| 2 | 87 |
| 3 | 40 |
| 4 | 7 |
| 5 | 9 |
| 6 | 7 |
| 7 | 0 |
| 8 | 1 |
| 9 | 1 |
| 10 | 0 |
| 11 | 0 |
| 12 | 0 |
| 13 | 0 |
| 14 | 0 |
| 15 | 0 |
| 16 | 1 |

**Table S2: *Staphylococcus aureus* specimen types in the study cohort.**

| Specimen Type | Number (Percentage) |
|---|---|
| Skin/Soft Tissue | 280 (33.7%) |
| Abscess | 162 (19.5%) |
| Pulmonary | 126 (15.1%) |
| Blood | 83 (10.0%) |
| Urine | 29 (3.5%) |
| Bone/Joint | 28 (3.4%) |
| Other | 4 (0.5%) |
| Uncategorized | 120 (14.4%) |
| Total | 832 |

**Resistance in the study cohort relative to 2022 institutional resistance**

Antibiograms at the Brigham and Women's Hospital and Massachusetts General Hospital are prepared using the isolate de-duplication methodology advised in the CLSI M39 document.[4] These guidelines recommend capturing only the first isolate from a given patient in a calendar year regardless of strain or body site. As described in the main text, we de-duplicated our isolates using a method intended to capture each unique episode of infection. **Table S3** compares our cohort to the institutional antibiograms when our cohort is de-duplicated via the per-infection method and the institutional antibiograms are de-duplicated via the CLSI method. **Table S4** compares rates of resistance when both our cohort and the institutional antibiograms are de-duplicated via the CLSI method. The CLSI method is more restrictive and identified only 623 unique specimens in our cohort compared to 832 specimens identified by the per-infection method described in the main text, reducing statistical power.

**Table S3: Resistance in our study cohort (de-duplicated via the per-infection method) relative to institutional resistance (de-duplicated by the CLSI method)**

| Antibiotic | Resistance in Study Cohort | Institutional Resistance | OR (95% CI, p-value) |
|---|---|---|---|
| **Tetracycline** | 123/829 (14.8%) | 244/2300 (10.6%) | 1.47 (1.15 – 1.86 , p = 0.002) |
| **Doxycycline** | 28/620 (4.5%) | 104/3477 (3.0%) | 1.53 (0.96 – 2.37, p = 0.06) |
| **Oxacillin** | 374/827 (45.2%) | 1492/5777 (25.8%) | 2.37 (2.04 – 2.76, p < 0.001) |
| **Trimethoprim-sulfamethoxazole** | 99/829 (11.9%) | 316/5777 (5.5%) | 3.24 (1.83 – 2.99, p < 0.001) |
| **Clindamycin** | 283/831 (34.1%) | 1573/5777 (27.2%) | 1.38 (1.18 – 1.61, p < 0.001) |

**Table S4: Resistance in the study cohort relative to institutional resistance in 2022 when both are de-duplicated using the CLSI method.**

| Antibiotic | Resistance in Study Cohort | Institutional Resistance | OR (95% CI, p-value) |
|---|---|---|---|
| **Tetracycline** | 81/620 (13.1%) | 244/2300 (10.6%) | 1.27 (0.96 – 1.67 , p = 0.10) |
| **Doxycycline** | 21/455 (4.6%) | 104/3477 (3.0%) | 1.57 (0.92 – 2.56, p = 0.09) |
| **Oxacillin** | 267/619 (43.1%) | 1492/5777 (25.8%) | 2.18 (1.83 – 2.59, p < 0.001) |
| **Trimethoprim-sulfamethoxazole** | 68/620 (11.0%) | 316/5777 (5.5%) | 2.12 (1.59 – 2.82, p < 0.001) |
| **Clindamycin** | 202/622 (32.5%) | 1573/5777 (27.2%) | 1.29 (1.07 – 1.54, p = 0.006) |

**References:**


1. R Core Team (2021). R: A language and environment for statistical computing. R Foundation for Statistical Computing, Vienna, Austria. URL https://www.R-project.org/.

2. Wickham H, Averick M, Bryan J, Chang W, McGowan LD, François R, Grolemund G, Hayes A, Henry L, Hester J, Kuhn M, Pedersen TL, Miller E, Bache SM, Müller K, Ooms J, Robinson D, Seidel DP, Spinu V, Takahashi K, Vaughan D, Wilke C, Woo K, Yutani H (2019). "Welcome to the tidyverse." _Journal of Open Source Software_, *4*(43), 1686. doi: 10.21105/joss.01686 (URL: https://doi.org/10.21105/joss.01686).

3. Long JA (2022). _jtools: Analysis and Presentation of Social Scientific Data_. R package version 2.2.0, <URL: https://cran.r-project.org/package=jtools>.

4. CLSI. Analysis and Presentation of Cumulative Antimicrobial Susceptibility Test Data; Approved Guideline -- Fourth Edition. CLSI document M39-A4. Wayne, PA. Clinical and Laboratory Standards Institute, 2014.
